# App usability, engagement, and postpartum weight retention: secondary analysis of the INTER-ACT randomized controlled trial

**DOI:** 10.64898/2025.12.09.25341940

**Authors:** Lisanne Duizer, Femke Geusens, Emma Geerits, Roland Devlieger, Annick Bogaerts

## Abstract

**Background:** Postpartum weight retention (PPWR) is a key contributor to long-term obesity risk and an important determinant of maternal health. Mobile health (mHealth) interventions may support postpartum change to a healthier lifestyle, but their effectiveness may depends on usability. This study examined whether perceived usability of the INTER-ACT app was associated with weight loss at 6 months postpartum and whether this association was mediated by app use frequency, motivational power, and implementation of lifestyle recommendations.

**Methods:** This secondary analysis used data from the intervention arm of the INTER-ACT randomized controlled trial (RCT), which provided postpartum lifestyle coaching and a supportive app to women with excessive gestational weight gain (EGWG). Participants (n=138) completed the process evaluation survey at 6 months postpartum, which included the System Usability Scale (SUS), perceived motivational power, and perceived implementation of lifestyle recommendations. Weight loss was calculated between 6 weeks and 6 months postpartum. Pearson correlations and parallel mediation analyses tested associations between usability, engagement-related variables, and weight loss, adjusting for maternal age, parity, and pre-pregnancy BMI.

**Results:** The INTER-ACT app demonstrated moderate usability (mean SUS=60.6). Usability was positively associated with app use frequency (B=.05, p<.001), perceived motivational power (B=.003, p<.001), and perceived implementation of lifestyle recommendations (B=.01, p=.036). However, none of these were associated with weight loss at 6 months postpartum. Usability showed no total, direct, or indirect effect on weight loss. Gestational weight gain (GWG) was the only significant predictor (B=0.24, p=0.003). Higher usability was also associated with more positive and fewer negative emotional responses (p<.05).

**Conclusion:** App usability was associated with engagement and positive emotional experience, but did not translate into reduced PPWR. GWG remained the main determinant of postpartum weight outcomes. Future interventions should prioritize preventing EGWG and complement this with emotionally supportive postpartum mHealth tools as part of a multi-faceted strategy.

## Introduction

Postpartum weight retention (PPWR), defined as the difference between pre-pregnancy weight and weight retained after childbirth, is a key contributor to long-term maternal obesity. Excessive postpartum weight retention (EPPWR; 5 kgs retained at 6 months postpartum) increases the risk of future cardiometabolic conditions such as type 2 diabetes and cardiovascular diseases, and is associated with poor outcomes in subsequent pregnancies, including gestational diabetes and hypertension (1–6). Psychosocially, persistent PPWR is linked to lower self-esteem, postpartum depression, and body dissatisfaction (7,8). Despite these risks, losing pregnancy weight remains challenging as around 75% of women do not return to their pre-pregnancy weight within the first postpartum year (9). Given these wide-ranging clinical and psychological implications, reducing PPWR is a key priority in postpartum care.

The postpartum period is characterized by profound physiological, emotional, and logistical challenges, including recovery from childbirth, sleep disruption, time scarcity, and the substantial responsibilities of caring for a newborn (10–12). These barriers complicate efforts to establish and maintain healthier behaviors. Effective interventions during this period must therefore be easily accessible, adaptable to fluctuating daily routines, and capable of supporting motivation over time (13). Considering these challenges, mobile health (mHealth) tools offer unique opportunities. They can provide flexible, on-demand, and personalized support and have the potential to reach women who are otherwise difficult to engage through traditional care models (14).

However, the effectiveness of mHealth interventions depends strongly on usability. According to the ISO 9241-11 framework, an international standard for evaluating human-system interaction, usability refers to how effectively, efficiently, and satisfactorily users can achieve intended goals within a specific context of use (15). This definition is particularly relevant for postpartum women, who frequently interact with apps under conditions of time pressure, fatigue, and emotional stress (16). Low usability can hinder task completion, evoke frustration, and ultimately contribute to disengagement. Conversely, well-designed apps may facilitate continued use, sustain motivation, and promote behavioral change (17,18). Previous research has also shown that evidence-based intervention apps often score lower on usability compared to commercial pregnancy apps, reducing their competitiveness given that many mothers prefer the more accessible commercial alternatives (19,20).

Usability alone, however, does not capture the broader experience of interacting with a digital tool. User experience (UX) is a multi-faceted concept that extends beyond usability and user interface design. While usability focuses on the effectiveness, efficiency, and ease of use of a system, UX encompasses broader experiential factors, including emotional responses, aesthetics, engagement, and perceived usefulness (15). UX is influenced not only by the characteristics of the product and the context in which it is used but also by users’ internal states, such as expectations, needs, motivations, and mood (21). For postpartum women, this means that an app is not only evaluated based on its functionality but also on how it makes them feel during moments of stress or vulnerability.

Furthermore, usability is intrinsically linked to the users’ emotional experience. A difficult-to-navigate app can evoke frustration and annoyance, whereas an easy-to-use app can foster feelings of competence and satisfaction (22). For this population, digital tools may serve either as a source of support or as an additional burden, making it crucial to understand these emotional responses when designing engaging and supportive interventions (23). Thus, UX extends beyond whether a task can be completed; it also reflects whether the experience is encouraging, reassuring, or overwhelming, which may influence users’ engagement and subsequent behavioral outcomes.

The quantity and organization of information, such as tips or recommendations, should be adjusted to align with users’ cognitive capacity so that the material remains manageable (24). Content perceived as too overwhelming, for example an excessive number of tips, can increase intrinsic and extraneous cognitive load, thereby diminishing perceived usability and potentially leading to disengagement. Conversely, too little information may fail to meet users’ needs, similarly undermining usability and trust in the system (25).

Although mHealth interventions are increasingly used to support postpartum women, no studies to our knowledge have directly examined how app usability affects behavioral implementation and PPWR outcomes. Evidence from other intervention fields suggests that app usability can significantly affect user engagement and behavioral change (26,27). However, it remains unclear whether these findings translate to postpartum women, who face unique challenges in the early months after childbirth. Emerging evidence suggests that more frequent app engagement is associated with healthier eating behaviors and weight loss among postpartum women (28,29).

Based on these insights, we hypothesize that usability is associated with behavioral outcomes. Specifically, if an app is perceived as easy to use and supportive, users may be more likely to improve their diet or physical activity, thereby reducing PPWR. In contrast, poor usability may hinder behavioral change regardless of content quality.

To address this gap, we examine the association between usability features of a postpartum mHealth tool and PPWR. The analysis is embedded within the INTER-ACT randomized controlled trial (RCT), which targets women with excessive gestational weight gain (EGWG), a group at elevated risk of PPWR (30). Within this framework, we examine the engagement pathways linking usability to postpartum outcomes. Three mediators are included in the model to clarify the mechanisms through which usability affects PPWR: app use frequency, perceived motivational power, and perceived implementation of lifestyle recommendations. In doing so, this study aims to provide actionable insights for designing and evaluating mHealth tools in maternal healthcare, with a focus on usability, user experience, emotional engagement, and effective content delivery.

## Materials and methods

### Study design and participants

This secondary analysis utilized data from the intervention arm of the INTER-ACT (INTERpregnAncy Coaching for a healthy fuTure) multi-center randomized controlled trial (RCT), conducted between 2017 and 2019 across 6 Flemish hospitals. The trial aimed to improve postpartum and interpregnancy health among women with EGWG (31,32).

Eligible participants were Dutch-speaking women aged 18 years or older who had delivered a singleton infant and exceeded the recommended guidelines of the Institute of Medicine (IOM) for gestational weight gain (GWG) (33). According to the IOM guidelines, recommended GWG ranges are 12.5-18 kg for women with underweight, 11.5-16 kg for women with normal weight, 7-11.5 kg for women with overweight, and 5-9 kg for women with obesity. Exclusion criteria were chronic illnesses requiring dietary restrictions or medication, psychiatric conditions, or contraindications for physical activity.

For this analysis, only women randomized to the intervention group who completed the process evaluation survey (covering user experience (UX)) and had available weight data at 6 weeks (baseline for app use) and 6 months postpartum were included. Women with missing survey or outcome data were excluded, resulting in a final sample of 138 participants (Fig 1). Baseline characteristics (pre-pregnancy weight, GWG, and weight loss at 6 weeks and 6 months postpartum) of the included sample did not differ from those of the full intervention group.

**Fig 1.**
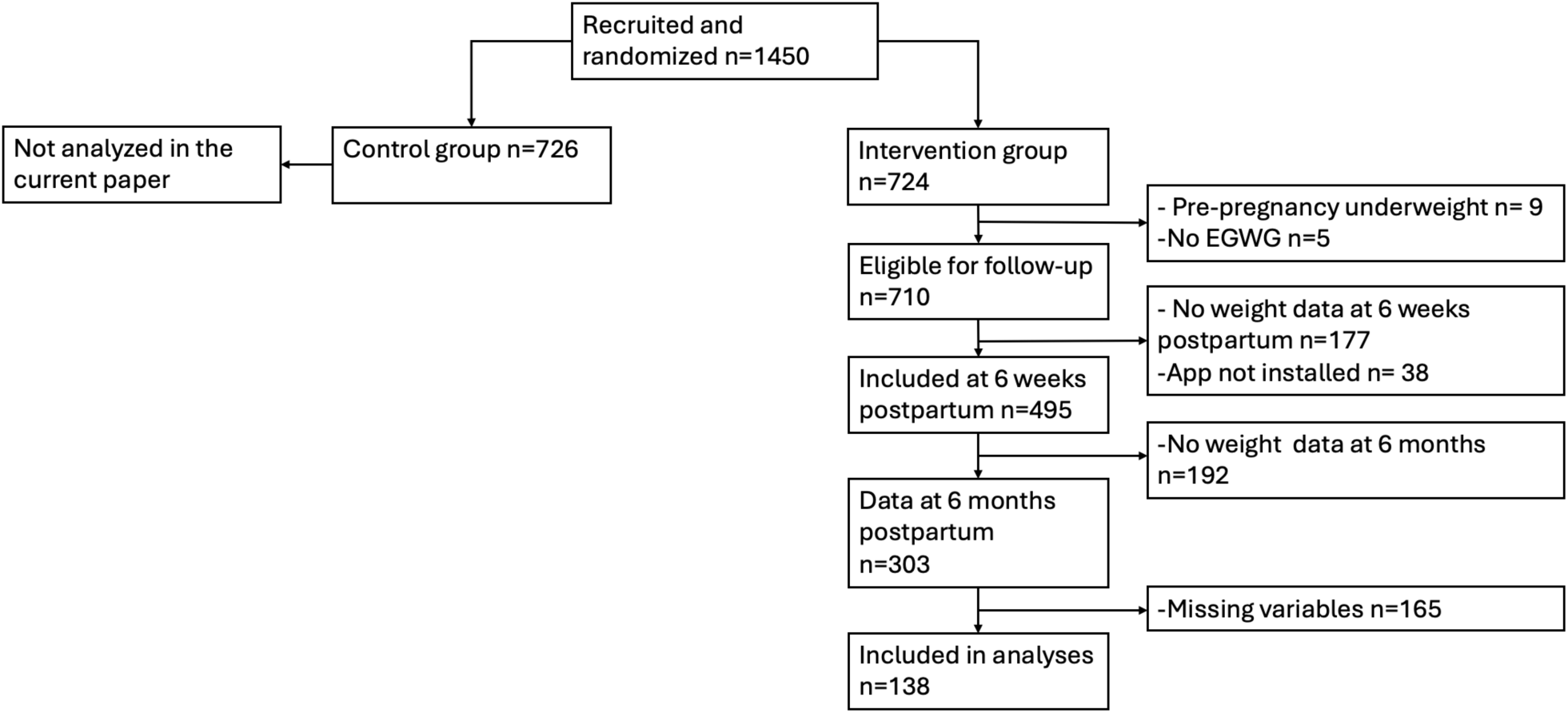
Flowchart of participant recruitment and final sample size.

### Intervention and app use

The INTER-ACT intervention consisted of 2 phases: a postpartum phase (6 weeks to 6 months postpartum) and an interpregnancy phase covering the preconception phase of a subsequent pregnancy. This study focuses exclusively on the postpartum phase.

Participants received 4 structured, face-to-face lifestyle coaching sessions at approximately 6, 8, and 12 weeks, and at 6 months postpartum. Coaching addressed 3 behavioral domains: nutrition (with emphasis on eating behavior), physical activity (including sedentary behavior reduction), and mental well-being.

In addition to coaching, participants received access to the INTER-ACT app, designed to support behavior change between sessions. The app provided personalized motivational messages, practical information, health-related advice, and individually tailored tips related to nutrition, activity, and emotional health. Users could define specific goals within the app, track progress, and record goal achievements.

To facilitate self-monitoring, participants were provided with a Bluetooth-connected weight scale and pedometer. Data from these devices were automatically synced with the app, allowing users to monitor their weight and physical activity levels and enabling coaches to provide personalized feedback during coaching sessions. In addition, participants also received periodic in-app questionnaires assessing lifestyle behaviors, well-being, engagement, and process evaluation. All app content was available exclusively in Dutch to ensure accessibility and ease of use for participants.

### Measures

#### Perceived app use frequency

Participants reported the number of days per week they used the app (0-7 days) on a scale in the evaluation survey.

#### Perceived app motivational power

Participants completed 4 self-developed Dutch items assessing perceived app motivational power. The items were: 1) the app motivated me to eat healthier, 2) the app motivated me to be more physically active, 3) the app helped me feel better about myself, and 4) the tracking of my weight changes on the app motivated me to adopt healthy lifestyle habits. Responses were provided on a 5-point Likert scale (1 = strongly disagree, 5= strongly agree).

#### Perceived implementation of lifestyle recommendations

Participants completed a set of 3 self-developed Dutch items assessing perceived implementation of lifestyle recommendations. The items were: 1) if I received a tip about nutrition, I applied it, 2) if I received a tip about physical activity, I applied it, and 3) if I received a tip about feeling good, I applied it.

Responses were provided on a 5-point Likert scale (1 = strongly disagree, 5= strongly agree). A Principal Component Analysis with Varimax rotation was conducted in SPSS to examine the underlying factor structure of the perceived implementation of lifestyle recommendations and perceived motivational power. The analysis revealed 2 distinct factors. The first factor, motivational app power, had an eigenvalue of 3.68 and explained 52.6% of the variance. The second factor, implementation of lifestyle recommendations, had an eigenvalue of 1.44 and explained 20.5% of the variance. Together, the 2 factors accounted for 73.1% of the total variance. Internal consistency was satisfactory for both factors (Cronbach’s α = 0.79 for motivational power; α = 0.89 for implementation). Mean scores were subsequently calculated separately for each factor and used in further analyses.

#### Perceived emotional responses

Participants were asked to indicate their perceived emotional responses to the following self-developed Dutch questions: 1) how did you feel while using the app, 2) how did you feel when reading tips about nutrition in the app, 3) how did you feel when reading tips about physical activity in the app, 4) how did you feel when reading tips about feeling good in the app, 5) how did you feel when seeing the number of steps you took in a day, 6) how did you feel when seeing your weight change in the app?

Participants were asked to indicate their emotional experience while using the app by selecting from a list of 10 emotions (happy, curious, persuaded, surprised, irritated, angry, sad, guilty, anxious, and insecure). For each question, the number of selected emotions was recorded, and a mean score was calculated across all 10 emotions. These mean scores were subsequently used in the analyses.

#### SUS score

Participants’ perception of app usability was assessed using the validated and widely used System Usability Scale (SUS) (34,35) (Supplementary 1). The SUS consists of 10 items, with responses recorded on a 5-point Likert scale (1 = strongly disagree, 5= strongly agree). Following standard scoring guidelines, responses to odd-numbered items were reverse-coded (36). Total SUS scores ranged from 0 to 100, with higher scores indicating better perceived usability. The SUS provides a global measure of usability, capturing 3 key dimensions: effectiveness (the extent to which users can achieve their goals accurately and completely), efficiency (the resources, such as time and effort, required to achieve those goals), and satisfaction (users’ comfort and positive attitudes toward using the app). By integrating these dimensions in a single score, the SUS score allows for rapid assessment of overall usability (35). Individual SUS scores were included in subsequent analyses to examine the relationship between perceived usability and other outcome measures.

#### Weight loss at 6 months postpartum

The primary outcome, PPWR, was calculated by subtracting the participant’s self-reported pre-pregnancy weight from the measured weight at 6 months postpartum, as recorded by trained study personnel. PPWR was analyzed both continuously (kg retained) and categorically, with substantial weight retention defined as ≥5 kilograms retained, a commonly used clinically meaningful indicator of long-term weight gain risk in postpartum research (2,37,38).

Because participants intitiated app use at 6 weeks postpartum, weight loss was defined as the difference between weight at 6 weeks and weight at 6 months postpartum to isolate the potential impact of app engagement.

#### Control variables

Maternal age (years), pre-pregnancy body mass index (BMI), and parity (primiparous vs. multiparous) were included as covariates in adjusted models. These variables were selected based on prior evidence linking them to either postpartum weight outcomes or intervention engagement (38,39).

#### Statistical analysis

All analyses were conducted using IBM SPSS Statistics version 29, with mediation models estimated using the PROCESS macro v5.0 (40). Descriptive statistics (means, SDs, percentages) were calculated for participant characteristics and key variables. Pearson correlations examined bivariate associations between usability (SUS score), perceived app use frequency, perceived app motivational power, and perceived implementation of lifestyle recommendations.

A parallel mediation model (PROCESS Model 4) was used to test whether the SUS scores were associated with weight loss at 6 months postpartum through 3 mediators, i.e. perceived motivational power, perceived implementation of lifestyle recommendations, and perceived app use frequency. The model was adjusted for pre-pregnancy BMI, maternal age, and parity. Bias-corrected 95% confidence intervals for indirect associations were estimated using 5000 bootstrap resamples, with associations considered significant when confidence intervals did not include zero. To examine whether app usability was associated with participants’ perceived emotional response, a multivariate general linear model (GLM) was conducted with SUS scores as the independent variable and the set of emotional response indicators as dependent variable.

## Results

### Participants characteristics

Descriptive statistics for the sample (N = 138) are presented in Table 1. Before pregnancy, participants weighed on average 71.16 kg (SD = 11.58). Weight loss averaged 11.01 kg (SD = 3.50) at 6 weeks postpartum, with an additional 2.69 kg (SD = 3.39) lost between 6 weeks at 6 months postpartum. Mean PPWR at 6 months postpartum was 4.08 kg (SD = 4.52), and 39.1% of participants exhibited EPPWR.

**Table 1.** Participant characteristics of the INTER-ACT intervention arm.

|  | Min | Max | Mean (SD) |
| --- | --- | --- | --- |
| **Weight (kg)** |  |  |  |
| Pre-pregnancy weight | 47.0 | 105.0 | 71.16 (11.58) |
| Weight loss at 6 weeks postpartum | -5.60<sup>a</sup> | 21.30 | 11.01 (3.50) |
| Weight loss at 6 months postpartum | -4.70<sup>a</sup> | 12.70 | 2.69 (3.39) |
| PPWR | -8.40<sup>a</sup> | 18.30 | 4.08 (4.52) |
| Total GWG | 10.0 | 33.7 | 17.78 (4.14) |
| Pre-pregnancy BMI | 18.87 | 37.11 | 25.54 (4.10) |
| **Demographics** |  |  |  |
| Age | 21 | 40 | 30.78 (3.85) |
| Children | 1 | 4 | 1.65 (0.79) |
| **App days per week** |  |  |  |
| Perceived app use frequency | 0.00 | 7.00 | 3.85 (1.91) |
| **Perceived usability** |  |  |  |
| SUS score | 15.00 | 100 | 60.60 (14.35) |
|  |  | N-% |  |
| Multiparous |  | 47.8% |  |
| Exclusively breastfeeding 6 weeks postpartum |  | 70.3% |  |
| EPPWR |  | 39.1% |  |
| **Emotional responses to app use** |  |  |  |
| Happiness |  | 38.3% |  |
| Irritation |  | 21.9% |  |
| Curiosity |  | 37.7% |  |
| Sadness |  | 5.6% |  |
| Guilty |  | 11.1% |  |
| Insecurity |  | 9.7% |  |
| Surprised |  | 8.2% |  |
| Angry |  | 2.8% |  |
| Persuaded |  | 15.9% |  |
| Anxiety |  | 0.5% |  |

288 **Note:** <sup>a</sup> Negative values indicate weight gain. SUS, System Usability Scale.

Total GWG averaged 17.78 kg (SD = 4.14), with participants exceeding the recommended IOM weight gain guidelines by an average of 4.59 kg (SD = 3.95). The mean pre-pregnancy BMI was 25.54 (SD = 4.10), placing most participants in the normal-weight to overweight range. Participants were, on average, 30.78 years old (SD = 3.85) and 47.8% were multiparous). At 6 weeks postpartum, 70.3% of participants exclusively breastfed the infant. Participants reported using the app on average 3.85 days per week (SD=1.91). The mean SUS score was 60.60 (SD=14.35), which is below the commonly used threshold of 68 indicating “good” usability. The most frequently reported emotional responses to the app were happiness (38.3%), curiosity (37.7%), and irritation (21.9%). Negative emotions such as sadness (5.6%), guilt (11.1%), insecurity (9.7%), and anger (2.8%) were reported less often. Feelings of being persuaded (15.9%) and surprised (8.2%) were also relatively low, while anxiety (0.5%) was scarcely reported. Furthermore, Pearson correlations indicated that the variables of interest were significantly associated with each other (Table 2).

**Table 2.**
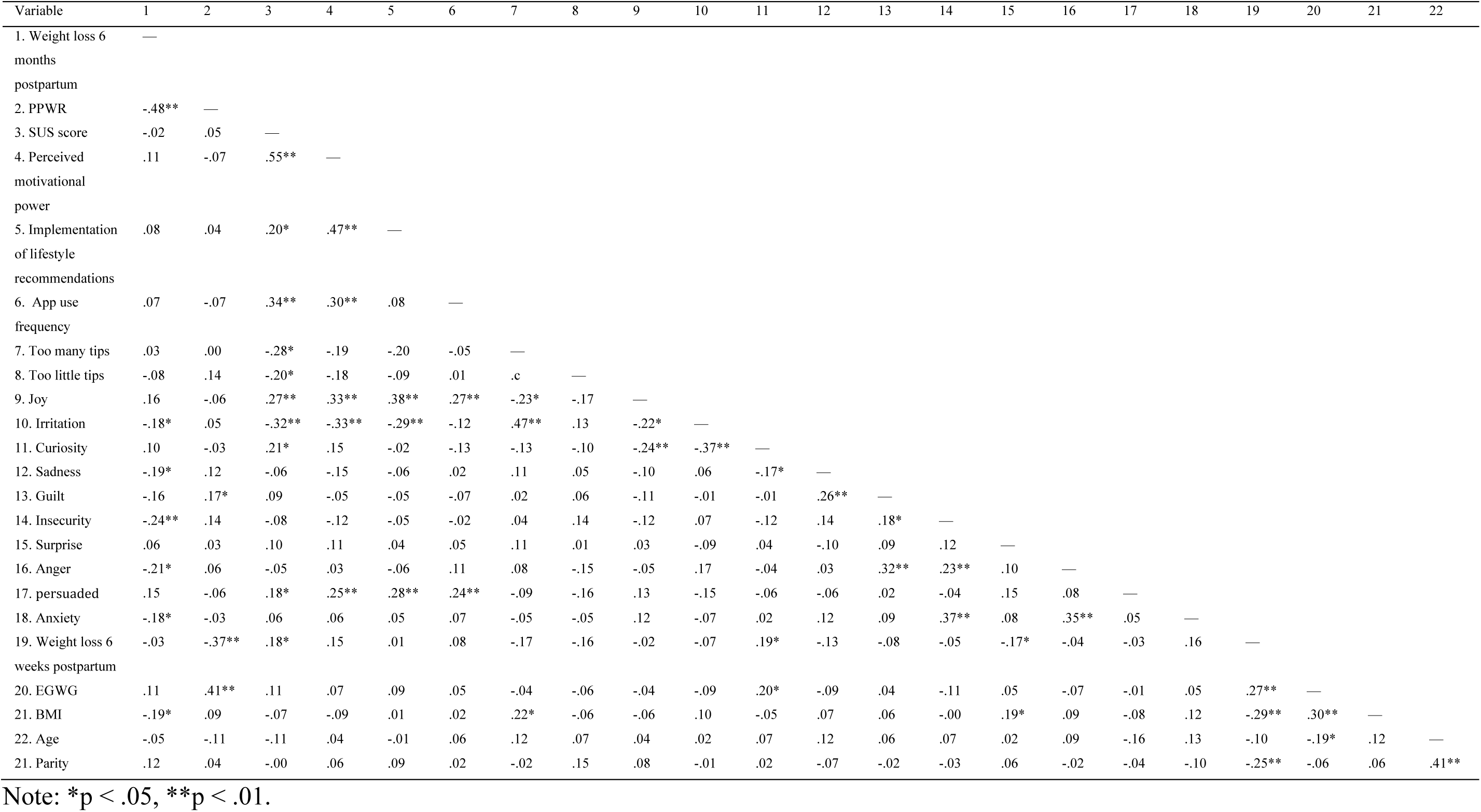
Pearson correlation matrix of weight loss, app usability, user experience, and emotional responses (N = 138).

| Variable | 1 | 2 | 3 | 4 | 5 | 6 | 7 | 8 | 9 | 10 | 11 | 12 | 13 | 14 | 15 | 16 | 17 | 18 | 19 | 20 | 21 | 22 |
| --- | --- | --- | --- | --- | --- | --- | --- | --- | --- | --- | --- | --- | --- | --- | --- | --- | --- | --- | --- | --- | --- | --- |
| 1. Weight loss 6 months postpartum | — |  |  |  |  |  |  |  |  |  |  |  |  |  |  |  |  |  |  |  |  |  |
| 2. PPWR | -.48** | — |  |  |  |  |  |  |  |  |  |  |  |  |  |  |  |  |  |  |  |  |
| 3. SUS score | -.02 | .05 | — |  |  |  |  |  |  |  |  |  |  |  |  |  |  |  |  |  |  |  |
| 4. Perceived motivational power | .11 | -.07 | .55** | — |  |  |  |  |  |  |  |  |  |  |  |  |  |  |  |  |  |  |
| 5. Implementation of lifestyle recommendations | .08 | .04 | .20* | .47** | — |  |  |  |  |  |  |  |  |  |  |  |  |  |  |  |  |  |
| 6. App use frequency | .07 | -.07 | .34** | .30** | .08 | — |  |  |  |  |  |  |  |  |  |  |  |  |  |  |  |  |
| 7. Too many tips | .03 | .00 | -.28* | -.19 | -.20 | -.05 | — |  |  |  |  |  |  |  |  |  |  |  |  |  |  |  |
| 8. Too little tips | -.08 | .14 | -.20* | -.18 | -.09 | .01 | .c | — |  |  |  |  |  |  |  |  |  |  |  |  |  |  |
| 9. Joy | .16 | -.06 | .27** | .33** | .38** | .27** | -.23* | -.17 | — |  |  |  |  |  |  |  |  |  |  |  |  |  |
| 10. Irritation | -.18* | .05 | -.32** | -.33** | -.29** | -.12 | .47** | .13 | -.22* | — |  |  |  |  |  |  |  |  |  |  |  |  |
| 11. Curiosity | .10 | -.03 | .21* | .15 | -.02 | -.13 | -.13 | -.10 | -.24** | -.37** | — |  |  |  |  |  |  |  |  |  |  |  |
| 12. Sadness | -.19* | .12 | -.06 | -.15 | -.06 | .02 | .11 | .05 | -.10 | .06 | -.17* | — |  |  |  |  |  |  |  |  |  |  |
| 13. Guilt | -.16 | .17* | .09 | -.05 | -.05 | -.07 | .02 | .06 | -.11 | -.01 | -.01 | .26** | — |  |  |  |  |  |  |  |  |  |
| 14. Insecurity | -.24** | .14 | -.08 | -.12 | -.05 | -.02 | .04 | .14 | -.12 | .07 | -.12 | .14 | .18* | — |  |  |  |  |  |  |  |  |
| 15. Surprise | .06 | .03 | .10 | .11 | .04 | .05 | .11 | .01 | .03 | -.09 | .04 | -.10 | .09 | .12 | — |  |  |  |  |  |  |  |
| 16. Anger | -.21* | .06 | -.05 | .03 | -.06 | .11 | .08 | -.15 | -.05 | .17 | -.04 | .03 | .32** | .23** | .10 | — |  |  |  |  |  |  |
| 17. persuaded | .15 | -.06 | .18* | .25** | .28** | .24** | -.09 | -.16 | .13 | -.15 | -.06 | -.06 | .02 | -.04 | .15 | .08 | — |  |  |  |  |  |
| 18. Anxiety | -.18* | -.03 | .06 | .06 | .05 | .07 | -.05 | -.05 | .12 | -.07 | .02 | .12 | .09 | .37** | .08 | .35** | .05 | — |  |  |  |  |
| 19. Weight loss 6 weeks postpartum | -.03 | -.37** | .18* | .15 | .01 | .08 | -.17 | -.16 | -.02 | -.07 | .19* | -.13 | -.08 | -.05 | -.17* | -.04 | -.03 | .16 | — |  |  |  |
| 20. EGWG | .11 | .41** | .11 | .07 | .09 | .05 | -.04 | -.06 | -.04 | -.09 | .20* | -.09 | .04 | -.11 | .05 | -.07 | -.01 | .05 | .27** | — |  |  |
| 21. BMI | -.19* | .09 | -.07 | -.09 | .01 | .02 | .22* | -.06 | -.06 | .10 | -.05 | .07 | .06 | -.00 | .19* | .09 | -.08 | .12 | -.29** | .30** | — |  |
| 22. Age | -.05 | -.11 | -.11 | .04 | -.01 | .06 | .12 | .07 | .04 | .02 | .07 | .12 | .06 | .07 | .02 | .09 | -.16 | .13 | -.10 | -.19* | .12 | — |
| 21. Parity | .12 | .04 | -.00 | .06 | .09 | .02 | -.02 | .15 | .08 | -.01 | .02 | -.07 | -.02 | -.03 | .06 | -.02 | -.04 | -.10 | -.25** | -.06 | .06 | .41** |

Note: \* $p < .05$ , \*\* $p < .01$ .

### App characteristics

#### SUS score

The SUS was used to assess overall app usability. The mean SUS score was 60.60 (SD=14.35), which indicates moderate usability and is slightly below the standard benchmark of 68 for “average” usability (Table 1).

### Mediation analysis: usability and weight loss at 6 months postpartum

A parallel mediation analysis (Model 4) tested whether SUS scores were associated with weight loss at 6 months postpartum through 3 mediators: perceived app use frequency, perceived motivational power, and perceived implementation of lifestyle recommendations (Table 3). SUS scores were significantly associated with more frequent app use (b= 0.05, p<0.001), greater perceived motivational power (b = 0.01, *p* < .036), and greater implementation of lifestyle recommendations (b=0.01, p= 0.036). Thus, better usability was associated with more frequent use, greater motivational impact, and higher perceived implementation of behavioral advice. However, none of the 3 mediators were significantly associated with weight loss at 6 months postpartum. Among the covariates, only GWG was positively associated with weight loss at 6 months postpartum (b=0.24, p=0.003).

**Table 3.**
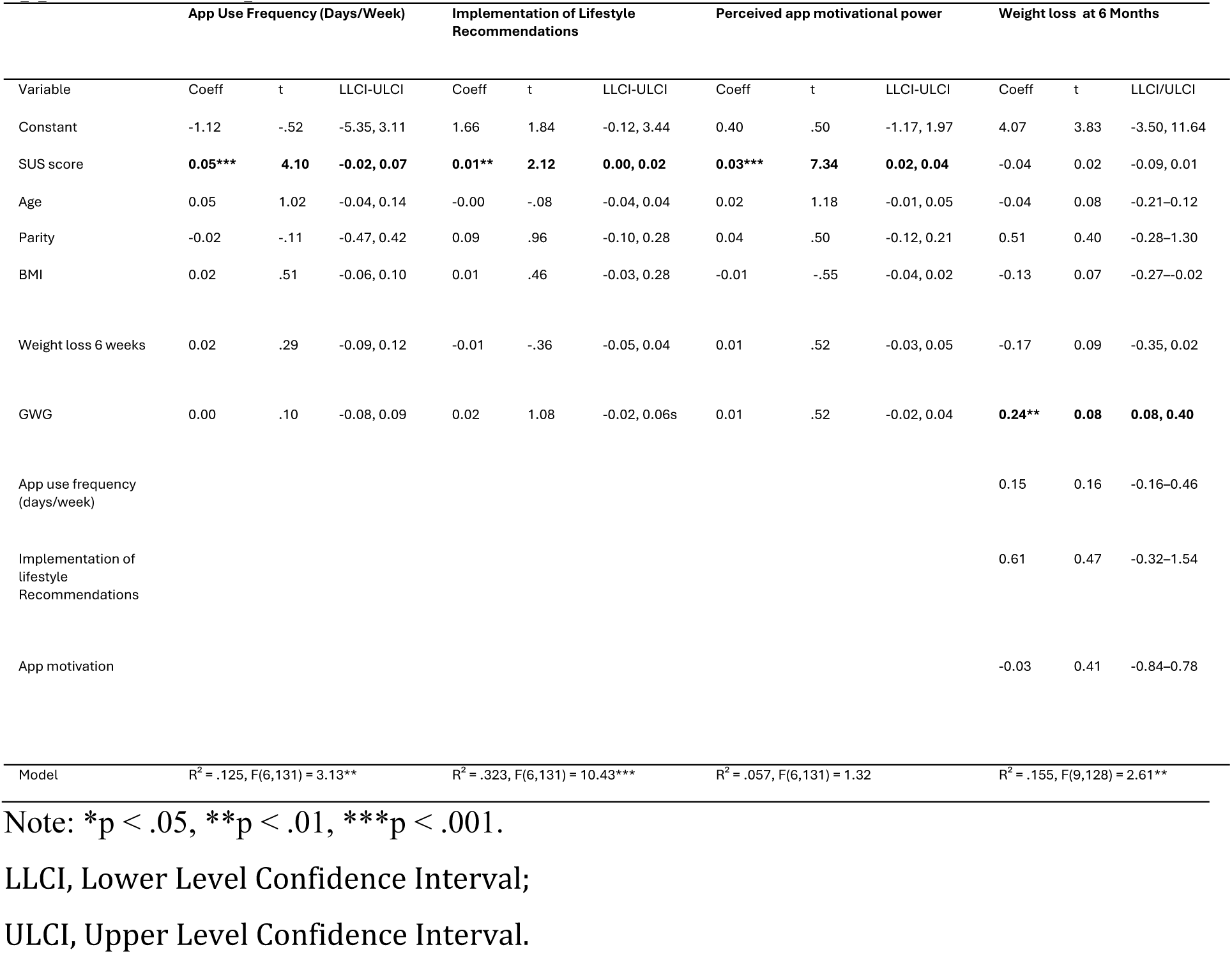
Mediation analyses of SUS Scores on weight loss at 6 months postpartum mediated through app use frequency, implementation of lifestyle recommendations, and perceived app motivational power.

Table 4 and Fig 2. summarize the total, direct, and indirect effects. Neither the total effect (b=-0.01, SE =0.02), nor the direct effect (b=-0.04, SE=0.02) of SUS score on weight loss at 6 months postpartum was significant. All indirect effects had confidence intervals that included zero, indicating no mediation.

**Table 4.** Overview of bootstrapped (5000 samples) standardized total, direct, and indirect effects.

| Effect type | Effect (B) | SE / BootSE | 95% CI |
| --- | --- | --- | --- |
| **Total effect** | -0.01 | 0.02 | [-0.05, 0.02] |
| **Direct effect** | -0.04 | 0.02 | [-0.09, 0.01] |
| **Indirect effect (Total)** | 0.03 | 0.01 | [-0.00, 0.05] |
| – via App use frequency | 0.01 | 0.01 | [-0.01, 0.02] |
| – via perceived motivational power | 0.02 | 0.01 | [-0.01, 0.05] |
| – via Implementation of lifestyle recommendation | 0.00 | 0.00 | [-0.01, 0.01] |

**Fig 2.**
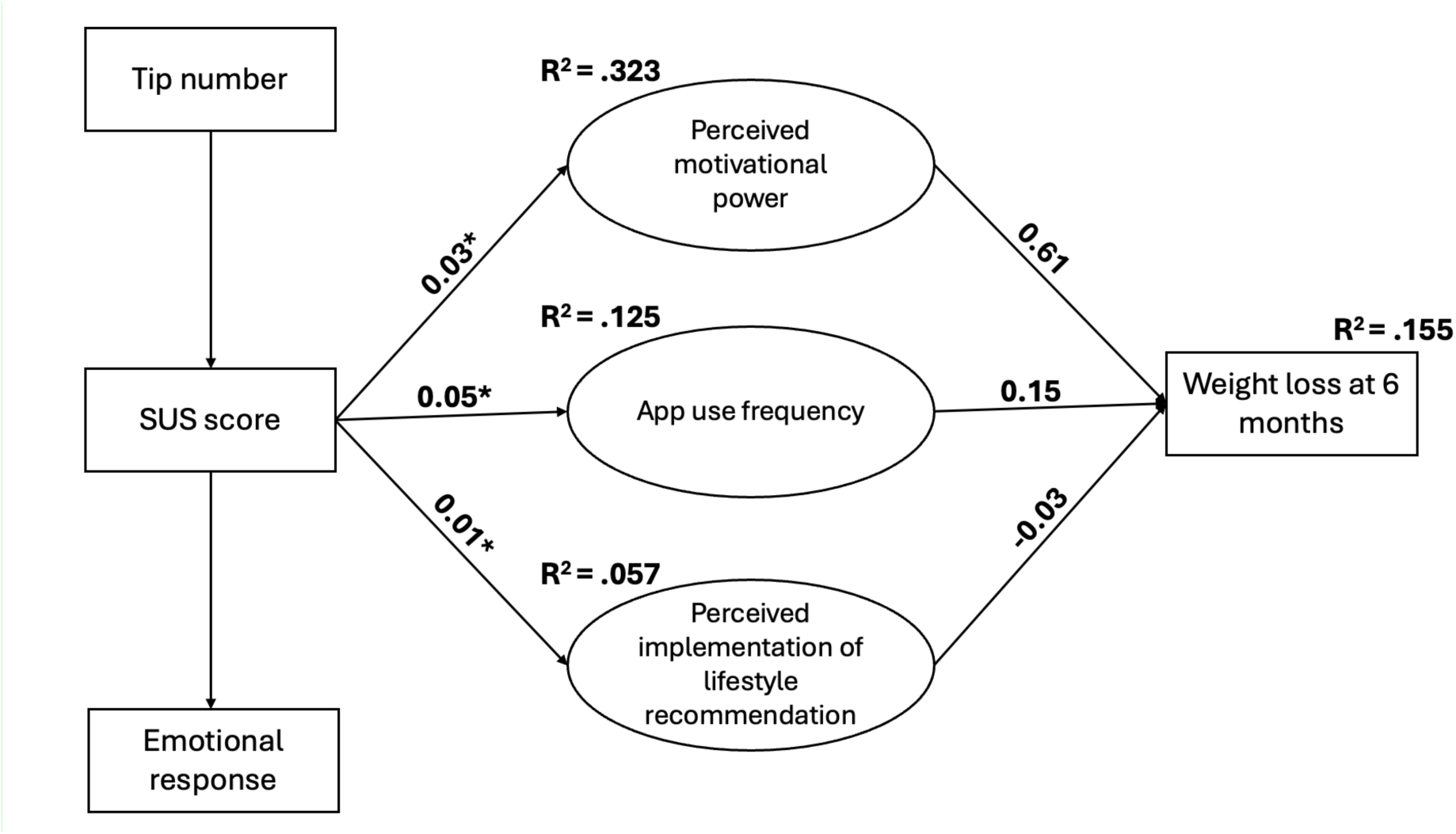
Proposed Conceptual Model of App-Based Intervention Effects.

#### Association between usability and emotional responses

A multivariate general linear model (GLM) evaluated whether SUS scores were associated with emotional responses to app use. The overall multivariate test was significant, Pillai’s Trace=.221, F(10, 123)=2.49, p<.001, partial η²=.221, indicating that perceived usability as measured by SUS was associated with the overall emotional response profile.

Follow-up univariate test showed that higher SUS scores were significantly associated with greater feelings of happiness (F(1,132) = 10.85, p=.001, partial η²=.076), less irritation (F(1,132)=14.21, p<.001), partial η²=.097), greater curiosity (F(1,132)=6.89, p=.010, partial η²=.050), and a modest increase in feelings of being persuaded (F(1,132)=3.91, p=.050, partial η²=.029) (Table 5). No significant associations emerged for sadness, guilt, insecurity, surprise, anger or anxiety (all ps >.20).

**Table 5.** Associations between SUS scores and individual emotional responses.

| Emotional Response | F(1,132) | p | Partial $\eta^2$ |
| --- | --- | --- | --- |
| **Happiness** | **10.85** | **.001** | **.076** |
| **Irritation** | **14.20** | **<.001** | **.097** |
| **Curiosity** | **6.89** | **.010** | **.050** |
| **Persuaded** | **3.91** | **.050** | **.029** |
| Sadness | 0.28 | .598 | .002 |
| Guilty | 1.30 | .256 | .010 |
| Insecurity | 0.72 | .397 | .005 |
| Surprised | 1.55 | .215 | .012 |
| Angry | 0.20 | .657 | .001 |
| Anxiety | 0.83 | .365 | .006 |

## Discussion

This study examined the relationship between perceived app usability, perceived app use frequency, perceived app motivational power, perceived implementation of lifestyle recommendations, and weight loss at 6 months postpartum among women using the INTER-ACT app. On average, women lost 11.01 kg (SD = 3.50) during the first 6 weeks postpartum and an additional 2.69 kg (SD = 3.39) between 6 weeks at 6 months postpartum, resulting in a mean PPWR of 4.08 kg (SD = 4.52). A substantial proportion (39%) showed EPPWR (≥5 kg), with some women even experiencing weight gain, underscoring the continued need for effective postpartum interventions to reduce PPWR.

In this context, evaluating the usability of the INTER-ACT app is crucial, as usability may influence adherence to lifestyle recommendations, perceived motivational power, and ultimately postpartum weight loss.

The mean SUS score of 60.60/100 suggests moderate usability, falling slightly below the commonly accepted benchmark of 68 for “average” usability (34,35,41). Although participants generally perceived the app as functional, these findings suggest there is potential to optimize the usability. Emotional responses were predominantly positive, with happiness and curiosity being reported most frequently, while negative emotions such as sadness, anger, and anxiety were rarely experienced. This pattern suggests that the app generally evoked favorable emotional reactions, which are increasingly recognized as important for sustained digital engagement (42). To further understand how these usability perceptions and affective responses relate to actual app behavior and adherence, correlation and mediation analyses were conducted.

The correlation and mediation analyses yielded two main insights.

First, perceived usability is significantly associated with perceived app use frequency, implementation of lifestyle recommendations, and motivational power. This aligns with prior literature suggesting that digital health tools with higher perceived usability promote sustained engagement and greater adherence (43). These results align with the foundational principle of human-computer interaction, which emphasizes usability as a core element (44). A well-designed, intuitive interface is a prerequisite for sustained engagement, as frustration or poor navigation can lead to disengagement regardless of potential health benefits (45). In addition, usability also shaped emotional responses: higher SUS scores were associated with greater happiness and curiosity and less irritation, suggesting that usability may act as both a technical and emotional determinant of UX.

Building on this, we also examined how the perceived balance of app content influenced usability ratings, highlighting another key factor for effective mHealth design.

The finding that perceiving the content as either too much or too little was associated with lower self-reported usability is particularly interesting for mHealth design. This supports a “Goldilocks” principle for digital health content: content should be neither overwhelming nor insufficient but calibrated so that users feel adequately supported without experiencing cognitive overload. Although this study did not directly test the role of content strategy or personalization, these factors could be important considerations for future mHealth design (46,47).

The second major insight is that this robust engagement pathway did not translate into significant weight loss. Despite clear associations between usability and engagement-related constructs, neither perceived app usability, perceived app use frequency, perceived motivational power, nor perceived implementation of lifestyle recommendations were significantly associated with weight loss at 6 months postpartum. This null finding is particularly important for the field of digital health interventions and highlights a key insight: perceived usability and perceived effectiveness do not necessarily associate with actual behavioral or clinical outcomes. Participants may feel that they are applying the recommendations, yet this perceived implementation did not result in meaningful weight loss. This raises the question of whether participants overestimate their adherence or whether the behavioral changes achieved were simply insufficient to produce measurable health benefits. Further research would benefit from combining self-report measures with objective markers of behavior (e.g., dietary logs, device-based activity tracking) to better capture the “dose” of behavioral change required to influence weight outcomes.

Beyond these factors, other explanations include the intensity and modality of the intervention. Although the INTER-ACT program combined the app with face-to-face coaching, the app itself may not have provided sufficiently individualized, adaptive, or intensive support needed to overcome the profound physiological, psychological, and lifestyle changes characteristic of the postpartum period.

The moderate usability level observed in this study may have further limited impact. While a SUS score of 60.6 appears adequate to support basic use, it also indicates notable usability shortcomings. It is plausible that minor frictions, such as slow loading times, confusing menus, or suboptimal notifications, may have disrupted the integration of app-based strategies into daily routines and reduced the consistency needed for weight loss. The app may have been “good enough” to use occasionally, but not “good enough” to support sustained behavioral change in an already demanding life phase.

Next, the results emphasize that postpartum weight regulation is shaped by multiple powerful determinants that extend beyond the scope of a single app. The only variable significantly associated with PPWR in our model was GWG, a well-established factor in the literature (30). This indicated that strong biological and pre-existing behavioral patterns (e.g. diet and exercise habits during pregnancy) may be more powerful determinants of postpartum weight than engagement with a postpartum mHealth tool. This raises the question of how much additional benefit can realistically be expected from lifestyle interventions initiated only after childbirth. Indeed, evidence from other interventions targeting PPWR suggests that, while some effects are observed, the overall impact is often modest and variable (48–50), highlighting the challenge of achieving substantial weight regulation through lifestyle changes alone. Unmeasured variables, such as sleep quality, stress, social support, mental health, and detailed dietary intake, likely contribute significantly as well (1,51,52).

At the same time, our findings show that higher usability was significantly associated with specific emotional responses. This points to a promising direction for future work: directly examining how discrete emotions, such as happiness, curiosity, irritation, and guilt shape, engagement trajectories over time. Emotional responses can influence how users perceive and interact with digital tools (42). Designing emotionally adaptive features, such as acknowledging user struggles or offering encouragement, may improve subjective experience, adherence, and ultimately health outcomes. Enhancing the usability of apps from moderate to good (SUS >70) can be achieved by minimizing sources of user irritation and customizing content delivery to better meet users’ emotional and practical needs, thereby improving overall satisfaction (53). Such strategies may also help address health equity concerns, while mobile apps provide accessible platforms for lifestyle support, their usability, emotional resonance, and perceived usefulness can differ widely across individuals, reflecting variations in digital health literacy, stress levels, or social support (50,51). Addressing these disparities in design and delivery will be essential to ensure that digital interventions equitably benefit diverse postpartum populations. This could be achieved through participatory or co-creation, where end-users are involved in the design and testing process to ensure cultural, emotional, and practical relevance. Furthermore, tailoring the app’s content and features to individual needs, such as offering varying degrees of guidance, feedback, or emotional tone, may help accommodate differences in stress, motivation, or support systems. Given the heterogeneity of postpartum experiences, a single “gold standard” design is unlikely; instead, adaptable, user-centered solutions are likely more effective.

Taken together, these findings suggest that while usability and positive user experience are essential foundations for engagement, they do not guarantee clinical effectiveness. The postpartum period presents unique and overwhelming challenges that may overshadow the impact of digital tools alone. Therefore, future interventions must integrate excellent, emotion-positive design into a more intensive, multi-faceted strategy, combining, for example, personalized digital coaching, social support, and structured behavioral modules, that directly addresses the complex biopsychosocial barriers faced by new mothers. To evaluate effectiveness, randomized controlled trials or longitudinal mixed-methods studies will be required, allowing assessment of both short and long-term outcomes while capturing user experience.

### Strengths and limitations

A major strength of this study is the use of a theory-driven mediation model to examine complex behavioral pathways, and the inclusion of a comprehensive set of factors, which provides a nuanced understanding of perceived effectiveness mechanisms in mHealth interventions. The multidimensional evaluation of usability (effectiveness, efficiency, and satisfaction) goes beyond typical usability studies that often rely on single-item or ad hoc measures. An additional strength is the involvement of a multidisciplinary team, combining expertise in psychology, behavioral science, nutrition, and clinical practice, which enhances the study’s comprehensiveness and applicability.

Another strength is the integration of psychological constructs (perceived motivational power), behavioral indicators (implementation of lifestyle recommendations), and clinical outcomes (weight loss at 6 months postpartum). Furthermore, by focusing on intermediate behavioral mechanisms such as perceived motivational power and implementation, this study addresses the “black box” often left unexplored in digital intervention research. Rather than focusing solely on end outcomes or app usage frequency, it investigates how specific components of UX may influence meaningful health behaviors. Finally, the combination of self-reported psychological measures with objective outcome data (e.g. weight change) strengthens the ecological validity of the findings.

Despite these strengths, several limitations must be acknowledged. First, the final analytic sample (N = 138) has limited power to detect small to moderate indirect effects. High attrition, largely due to missing data and participant dropout, may have introduced bias. It is possible that women who completed the follow-up were more motivated, satisfied with the app, or engaged in their health, which could skew results and limit generalizability. However, comparison of baseline characteristics between women included in the current analysis and the overall intervention group showed no significant differences, suggesting that the analytic sample was generally representative of the original intervention group.

Second, the sample consisted exclusively of Dutch-speaking women who had experienced EGWG, which may limit the generalizability of the findings to more diverse populations or those with different perinatal profiles.

Further research with larger and more diverse samples is needed to confirm whether the observed patterns represent true effects or reflects limited statistical power. Larger studies may also allow exploration of subgroup differences, identifying which intervention components (e.g. education, personalized feedback, social support, active monitoring) are most effective, and determining how these elements can be combined with usability and UX insight to improve both engagement and clinical outcomes. This could help identify for whom mHealth interventions are most effective and which components drive lifestyle changes

Another limitation is that some key measures were self-reported, including pre-pregnancy weight and BMI, which may introduce recall bias and affect data reliability. Lastly, the duration of the follow-up may have been insufficient to observe long-term effects of behavioral change on weight outcomes.

Future studies may benefit from mixed-method approaches that explore users’ subjective experiences in greater depth. Interviews or open-ended surveys could provide valuable insight into why some users remain engaged despite low satisfaction, or what design features postpartum users find most empowering or discouraging.

To enhance real-time insight into user engagement and behavior, future research could benefit from integrating ecological momentary assessment (EMA) or experience sampling methods (ESM), which would allow tracking of app use and emotional responses in daily life. Lastly, the study focused on outcomes within the first 6 months postpartum. While this is a crucial period for behavioral and physical adjustments, it may not fully capture the long-term sustainability of user engagement and weight-related changes. Furthermore, weight loss at 6 months postpartum was calculated as the difference between body weight at 6 weeks and 6 months postpartum, given that the intervention started at 6 weeks. This means that many women may have already experienced substantial weight loss, 11.01 kg (SD = 3.50), during the first 6 weeks, a period in which weight loss is often more pronounced due to natural postpartum recovery. As a result, the additional weight to be lost after 6 weeks may have been more limited, potentially reducing the need for or perceived benefit of the intervention. Consequently, women may have disengaged from the app, which could have contributed to slower weight loss during the later postpartum period.

## Conclusion

While app usability is associated with perceived app use frequency, perceived motivational power, and perceived implementation of lifestyle recommendations, it is not associated with reduced weight loss at 6 months postpartum. Furthermore, usability also shapes emotional responses: higher usability scores were associated with significantly greater feelings of happiness and curiosity, and less irritation. However, these factors did not translate into significant weight loss. The strongest factor associated with PPWR remained GWG, highlighting that the profound biological and contextual challenges of pregnancy can outweigh the effect of digital support in the postpartum period. Future research should therefore not only aim to identify which usability and behavioral components enhance engagement after childbirth but also focus on strategies to optimize GWG during pregnancy to prevent postpartum weight retention.

## Data Availability

All relevant data are within the manuscript and its Supporting Information files.

## Completing interests

The authors have no conflict of interest to report

## Ethical approval

This study was approved by the Clinical Trial Center/Ethical Committee UZ Leuven (protocol code B322201730956/S59889).

## Acknowledgments

We are grateful to the study midwives for their valuable assistance with participant recruitment. We also acknowledge the financial and logistical support provided by FWO Vlaanderen, Rotary Foundation Limburg, Horizon Europe, Agentschap kind en gezin/opgroeien (Flemish organization offering home-based nursing and midwifery services).

RD is holder of a personal BOF-FKO grant from KULeuven (2025–2030)

## Acknowledgments

Conceptualization: Femke Geusens, Annick Bogaerts, Lisanne Duizer.

Data curation: Hanne Van Uytsel, Lieveke Ameye, Margriet Bijlholt.

Formal analysis: Lisanne Duizer.

Funding acquisition: Annick Bogaerts, Roland Devlieger.

Investigation: Annick Bogaerts, Roland Devlieger.

Methodology: Annick Bogaerts, Roland Devlieger.

Project administration: Annick Bogaerts.

Resources: FWO, MSCA-JD.

Software: Lisanne Duizer.

Supervision: Annick Bogaerts, Femke Geusens.

Validation: Annick Bogaerts, Roland Devlieger.

Visualization: Lisanne Duizer.

Writing–original draft: Lisanne Duizer.

Writing–review & editing: Femke Geusens, Roland Devlieger, Annick Bogaerts, Emma Geerits.

## Supporting information

### S1 File. System Usability Score (SUS) questionnaire

1. I think that I would like to use this system frequently.
2. I found the system unnecessarily complex.
3. I thought the system was easy to use.
4. I think that I would need the support of a technical person to be able to use this system.
5. I found the various functions in this system were well integrated.
6. I thought there was too much inconsistency in this system.
7. I would imagine that most people would learn to use this system very quickly.
8. I found the system very cumbersome to use.
9. I felt very confident using the system.
10. I needed to learn a lot of things before I could get going with this system.

## Reference

1. Makama M, Skouteris H, Moran LJ, Lim S. Reducing Postpartum Weight Retention: A Review of the Implementation Challenges of Postpartum Lifestyle Interventions. J Clin Med. 2021 Apr 27;10(9):1891.

2. Fadzil F, Shamsuddin K, Wan Puteh SE, Mohd Tamil A, Ahmad S, Abdul Hayi NS, et al. Predictors of postpartum weight retention among urban Malaysian mothers: A prospective cohort study. Obes Res Clin Pract. 2018 Nov;12(6):493–9.

3. Kew S, Ye C, Hanley AJ, Connelly PW, Sermer M, Zinman B, et al. Cardiometabolic Implications of Postpartum Weight Changes in the First Year After Delivery. Diabetes Care. 2014 July 1;37(7):1998–2006.

4. Bogaerts A, Van Den Bergh BRH, Ameye L, Witters I, Martens E, Timmerman D, et al. Interpregnancy Weight Change and Risk for Adverse Perinatal Outcome. Obstet Gynecol. 2013 Nov;122(5):999–1009.

5. Liu J, Song G, Meng T, Zhao G, Guo S. Weight retention at six weeks postpartum and the risk of gestational diabetes mellitus in a second pregnancy. BMC Pregnancy Childbirth. 2019 Dec;19(1):272.

6. Kirkegaard H, Bliddal M, Støvring H, Rasmussen KM, Gunderson EP, Køber L, et al. Maternal weight change from prepregnancy to 18 months postpartum and subsequent risk of hypertension and cardiovascular disease in Danish women: A cohort study. Wareham NJ, editor. PLOS Med. 2021 Apr 2;18(4):e1003486.

7. Collings R, Hill B, Skouteris H. The influence of psychological factors on postpartum weight retention 12 months post-birth. J Reprod Infant Psychol. 2018 Mar 15;36(2):177–91.

8. Bazzazian S, Riazi H, Vafa M, Mahmoodi Z, Nasiri M, Mokhtaryan-Gilani T, et al. The relationship between depression, stress, anxiety, and postpartum weight retention: A systematic review. J Educ Health Promot. 2021 Jan;10(1):230.

9. Endres LK, Straub H, McKinney C, Plunkett B, Minkovitz CS, Schetter CD, et al. Postpartum Weight Retention Risk Factors and Relationship to Obesity at 1 Year. Obstet Gynecol. 2015 Jan;125(1):144–52.

10. Van Der Pligt P, Willcox J, Hesketh KD, Ball K, Wilkinson S, Crawford D, et al. Systematic review of lifestyle interventions to limit postpartum weight retention: implications for future opportunities to prevent maternal overweight and obesity following childbirth. Obes Rev. 2013 Oct;14(10):792–805.

11. Navigating the Postpartum Period: Hormonal Changes and Essential Care for Women. In: Postpartum Period for Mother and Newborn [Working Title] [Internet]. IntechOpen; 2025 [cited 2025 July 25]. Available from: https://www.intechopen.com/online-first/1207066

12. Hill B, McPhie S, Moran LJ, Harrison P, Huang TTK, Teede H, et al. Lifestyle intervention to prevent obesity during pregnancy: Implications and recommendations for research and implementation. Midwifery. 2017 June;49:13–8.

13. Lim S, Lang S, Savaglio M, Skouteris H, Moran LJ. Intervention Strategies to Address Barriers and Facilitators to a Healthy Lifestyle Using the Behaviour Change Wheel: A Qualitative Analysis of the Perspectives of Postpartum Women. Nutrients. 2024 Apr 3;16(7):1046.

14. Evans K, Rennick-Egglestone S, Cox S, Kuipers Y, Spiby H. Remotely Delivered Interventions to Support Women With Symptoms of Anxiety in Pregnancy: Mixed Methods Systematic Review and Meta-analysis. J Med Internet Res. 2022 Feb 15;24(2):e28093.

15. Dietlein CS, Bock OL. Development of a usability scale based on the three ISO 9241-11 categories “effectiveness,” “efficacy” and “satisfaction”: a technical note. Accreditation Qual Assur. 2019 June;24(3):181–9.

16. Jacob C, Sezgin E, Sanchez-Vazquez A, Ivory C. Sociotechnical Factors Affecting Patients’ Adoption of Mobile Health Tools: Systematic Literature Review and Narrative Synthesis. JMIR MHealth UHealth. 2022 May 5;10(5):e36284.

17. Perski O, Blandford A, West R, Michie S. Conceptualising engagement with digital behaviour change interventions: a systematic review using principles from critical interpretive synthesis. Transl Behav Med. 2017 June;7(2):254–67.

18. McKay FH, Wright A, Shill J, Stephens H, Uccellini M. Using Health and Well-Being Apps for Behavior Change: A Systematic Search and Rating of Apps. JMIR MHealth UHealth. 2019 July 4;7(7):e11926.

19. Tucker L, Villagomez AC, Krishnamurti T. Comprehensively addressing postpartum maternal health: a content and image review of commercially available mobile health apps. BMC Pregnancy Childbirth. 2021 Dec;21(1):311.

20. Tucker L, Villagomez AC, Krishnamurti T. Comprehensively addressing postpartum maternal health: a content and image review of commercially available mobile health apps. BMC Pregnancy Childbirth. 2021 Dec;21(1):311.

21. Hellweger S, Wang X. What is User Experience Really: towards a UX Conceptual Framework.

22. Saariluoma P, Jokinen JPP. Emotional Dimensions of User Experience: A User Psychological Analysis. Int J Hum-Comput Interact. 2014 Apr 3;30(4):303–20.

23. Hanach N, Saqan R, Radwan H, Baniissa W, De Vries N. Perceived Experiences and Needs of Digital Resources Among Postpartum Women in the United Arab Emirates: Qualitative Focus Group Study. J Med Internet Res. 2024 Dec 16;26:e53720.

24. Kennedy MJ, Romig JE. Cognitive Load Theory: An Applied Reintroduction for Special and General Educators. Teach Except Child. 2024 July;56(6):440–51.

25. Clarke MA, Schuetzler RM, Windle JR, Pachunka E, Fruhling A. Usability and cognitive load in the design of a personal health record. Health Policy Technol. 2020 June;9(2):218–24.

26. Lin YP, Lee KC, Ma WF, Syu BS, Liao WC, Yang HT, et al. A mobile technology-based tailored health promotion program for sedentary employees: development and usability study. BMC Public Health. 2025 Apr 17;25(1):1452.

27. Assessing the Impact of Usability from Evaluating Mobile Health Applications. Int J Eng Technol Inform. 2024 Feb 26;

28. Geusens F, Van Uytsel H, Ameye L, Devlieger R, Jacquemyn Y, Van Holsbeke C, et al. The impact of self-monitoring physical and mental health via an mHealth application on postpartum weight retention: Data from the INTER-ACT RCT. Health Promot Perspect. 2024 Mar 14;14(1):44–52.

29. Xiaocui H, Shengyao Y, Samsudin N, Kuan L, Xuefen L. Empowering postpartum women: the role of mHealth apps in promoting mental health and obesity prevention. BMC Womens Health. 2025 July 16;25(1):351.

30. He X, Hu C, Chen L, Wang Q, Qin F. The association between gestational weight gain and substantial weight retention 1-year postpartum. Arch Gynecol Obstet. 2014 Sept;290(3):493–9.

31. Bogaerts A, Van Uytsel H, Ameye L, Winter Shafran Y, Jacquemyn Y, Van Holsbeke C, et al. Interpregnancy and pregnancy lifestyle intervention (INTER-ACT): a randomized controlled trial. Am J Obstet Gynecol. 2025 Dec;233(6):668.e1–668.e13.

32. Bogaerts A, Ameye L, Bijlholt M, Amuli K, Heynickx D, Devlieger R. INTER-ACT: prevention of pregnancy complications through an e-health driven interpregnancy lifestyle intervention – study protocol of a multicentre randomised controlled trial. BMC Pregnancy Childbirth. 2017 Dec;17(1):154.

33. Rasmussen KM, Yaktine AL, Institute of Medicine (U.S.), editors. Weight gain during pregnancy: reexamining the guidelines. Washington, D. C: National Academies Press; 2009. 1 p.

34. Ensink CJ, Keijsers NLW, Groen BE. Translation and validation of the System Usability Scale to a Dutch version: D-SUS. Disabil Rehabil. 2024 Jan 16;46(2):395–400.

35. Brooke J. SUS - A quick and dirty usability scale.

36. Hyzy M, Bond R, Mulvenna M, Bai L, Dix A, Leigh S, et al. System Usability Scale Benchmarking for Digital Health Apps: Meta-analysis. JMIR MHealth UHealth. 2022 Aug 18;10(8):e37290.

37. Althuizen E, Van Poppel MN, De Vries JH, Seidell JC, Van Mechelen W. Postpartum behaviour as predictor of weight change from before pregnancy to one year postpartum. BMC Public Health. 2011 Dec;11(1):165.

38. Dol J, Richardson B, Murphy GT, Aston M, McMillan D, Campbell-Yeo M. Impact of mobile health interventions during the perinatal period on maternal psychosocial outcomes: a systematic review. JBI Evid Synth. 2020 Jan;18(1):30–55.

39. Archuleta J, Chao SM. Maternal Characteristics that Impact Postpartum Weight Retention: Results from the 2016 Los Angeles Mommy and Baby (LAMB) Follow-Up Study. Matern Child Health J. 2021 Jan;25(1):151–61.

40. Clement LM, Bradley-Garcia M. A Step-By-Step Tutorial for Performing a Moderated Mediation Analysis using PROCESS. Quant Methods Psychol. 2022 Oct 1;18(3):258–71.

41. Grier RA, Bangor A, Kortum P, Peres SC. The System Usability Scale: Beyond Standard Usability Testing. Proc Hum Factors Ergon Soc Annu Meet. 2013 Sept;57(1):187–91.

42. Szinay D, Perski O, Jones A, Chadborn T, Brown J, Naughton F. Perceptions of Factors Influencing Engagement With Health and Well-being Apps in the United Kingdom: Qualitative Interview Study. JMIR MHealth UHealth. 2021 Dec 16;9(12):e29098.

43. Hyzy M, Bond R, Mulvenna M, Bai L, Dix A, Leigh S, et al. System Usability Scale Benchmarking for Digital Health Apps: Meta-analysis. JMIR MHealth UHealth. 2022 Aug 18;10(8):e37290.

44. Dix A. Human–computer interaction, foundations and new paradigms. J Vis Lang Comput. 2017 Oct;42:122–34.

45. O’Brien HL, Roll I, Kampen A, Davoudi N. Rethinking (Dis)engagement in human-computer interaction. Comput Hum Behav. 2022 Mar;128:107109.

46. Nelson LA, Spieker AJ, LeStourgeon LM, Greevy Jr RA, Molli S, Roddy MK, et al. The Goldilocks Dilemma on Balancing User Response and Reflection in mHealth Interventions: Observational Study. JMIR MHealth UHealth. 2024 Jan 19;12:e47632–e47632.

47. IISE Transactions on Occupational Ergonomics and Human Factors. IISE Trans Occup Ergon Hum Factors. 2017 Oct 2;5(3–4):ebi-ebi.

48. Falciglia G, Piazza J, Ollberding NJ, Spiess L, Morrow A. A Theory-Based Dietary Intervention for Overweight, Postpartum Mothers and Their Children Improves Maternal Vegetable Intake. Open J Obstet Gynecol. 2017;07(07):679–92.

49. Wilkinson SA, Van Der Pligt P, Gibbons KS, McIntyre HD. Trial for R educing W eight R etention in N ew M ums: a randomised controlled trial evaluating a low intensity, postpartum weight management programme. J Hum Nutr Diet. 2015 Jan;28(s1):15–28.

50. Gilmore LA, Klempel MC, Martin CK, Myers CA, Burton JH, Sutton EF, et al. Personalized Mobile Health Intervention for Health and Weight Loss in Postpartum Women Receiving Women, Infants, and Children Benefit: A Randomized Controlled Pilot Study. J Womens Health. 2017 July;26(7):719–27.

51. Gunderson EP, Rifas-Shiman SL, Oken E, Rich-Edwards JW, Kleinman KP, Taveras EM, et al. Association of Fewer Hours of Sleep at 6 Months Postpartum with Substantial Weight Retention at 1 Year Postpartum. Am J Epidemiol. 2007 Oct 17;167(2):178–87.

52. Whitaker K, Young-Hyman D, Vernon M, Wilcox S. Maternal Stress Predicts Postpartum Weight Retention. Matern Child Health J. 2014 Nov;18(9):2209–17.

53. Oyebode O, Alqahtani F, Orji R. Using Machine Learning and Thematic Analysis Methods to Evaluate Mental Health Apps Based on User Reviews. IEEE Access. 2020;8:111141–58.

